# Effects of Chinese strategies for controlling the diffusion and deterioration of novel coronavirus–infected pneumonia in China

**DOI:** 10.1101/2020.03.10.20032755

**Authors:** Xiaoqiang Wang, Weitian Tian, Xin Lv, Yumiao Shi, Xiaoxin Zhou, Weifeng Yu, Diansan Su, Jie Tian

## Abstract

**Background:** In December 2019, an outbreak of new type of coronavirus named COVID-19 occurred in Wuhan, Hubei Province, China. In a very short time, this virus spread rapidly over China, greatly threatening public health and economic development. The Chinese government acted quickly and implemented a series of strategies to prevent diffusion of this disease. We therefore sought to evaluate the effects of these Chinese strategies for controlling the spread of COVID-19.

**Methods:** From the data of cumulative confirmed cases from provincial Health Commission websites of China, we performed model fitting and calculated the growth speed of cumulative confirmed patients. We further analyzed the time when this growth speed, the rate of the number of new cases, reached its maximum (Speed_max_). Comparing different times to Speed_max_ of different areas in China, we calculated the dates at which the growth speed began to decline in different areas. Also, The number of plateaus were analyzed.

**Results:** The quartic model showed the best fit. For almost all areas in mainland China, the speed of infections reached Speed_max_ and began to decline within 14 days; exceptions were Hebei, Heilongjiang, Hainan, Guizhou, and Hubei. The number of plateaus was significantly correlated with the emigration index. However, the distance from other areas to Hubei and the number of plateaus had little influence on when a province or area arrived at Speed_max_. Once strict intervention strategies were implemented, diffusion and deterioration of COVID-19 were inhibited quickly and effectively over China.

**Conclusion:** Our study suggests that Chinese strategies are highly effective on controlling the diffusion and deterioration of the novel coronavirus–infected pneumonia. These strategies supply experience and guidelines for other countries to control the COVID-19 epidemic.

## Introduction

On December 8, 2019, the first patient with atypical pneumonia of unknown origin was reported by the government of Wuhan, Hubei, China ^[1]^. Since then, the outbreak in Wuhan became an epidemic, rapidly spreading through China and other countries. On January 1, 2020, a new type of coronavirus was isolated and finally named COVID-19 by the World Health Organization on February 11, 2020 ^[2, 3]^. According to reports ^[4]^, by March 1, 2020, more than 60 countries or areas had reported the confirmed patients with COVID-19, and more than 80,000 patients had been confirmed with COVID-19 worldwide. This has serious implications for both public health and development in China and the world.

To control the diffusion and deterioration of the epidemic of COVID-19 in China, the Chinese government implemented series of strict and unprecedented intervention strategies ^[5, 6]^. For example, on January 21, 2020, COVID-19 was classed as a Class B infectious disease and was controlled as a Class A infectious disease; on January 23, 2020, Wuhan was quarantined to limit the diffusion of patients with COVID-19. All other provinces and areas in China also began to control public gatherings and the migration of people from January 21, 2020. These mandatory strategies carried out by the government showed significant effects on containing the contagion of COVID-19, and the number of new confirmed cases remarkably decreased at the end of February 2020.

To estimate optimally the effects of Chinese strategies on controlling the diffusion and deterioration of COVID-19 in China, we performed model fitting and calculated the time at which the growth speed of the cumulative number of confirmed patients reached the maximum.

## Methods

### Data sources

For COVID-19 data, we collected the number of cumulative confirmed patients from January 21, 2020 to February 23, 2020 because almost all areas in China began to report confirmed cases from January 21, 2020, and the growth speed reached zero around most areas except Hubei before February 23, 2020. The number of cumulative confirmed patients with COVID-19 in different provinces or areas were acquired from the respective Health Commission websites of China. For COVID-19 data, areas of Hong Kong, Macao, and Taiwan were not analyzed in this study. The emigration index in mainland China was obtained from the Baidu Qianxi website (http://qianxi.baidu.com/).

### Model fitting for known data

In this study, we tried multiple curve models to fit known data and times. We used the following models to attempt a good fit of the data: exponential, logarithmic, power function, compound, growth, inverse, logistic, linear, quadratic, cubic, and quartic. Of these, the quartic model showed the best fit, and almost all correlation coefficients reached R^2^ > 0.99 for fitted curves in different areas. With time defined as *x* and January 21, 2020 was recorded as the time “1,” the number of cumulative confirmed patients with COVID-19 (*y*) and time (*x*) were expressed as the following quartic function:

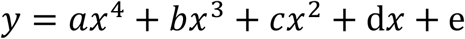

### The growth speed calculation for fitted curves

According to fitted curves modeled by known data, we found that almost all fitted curves showed an “S” form. This suggested that the growth speed of cumulative confirmed cases increased first and gradually decreased after reaching the maximum. Therefore, we analyzed the continuous change of the growth speed and obtained the time when the growth speed reached the maximum (Speed_max_) by drawing the derivative function of the fitted curves. When the time was defined as *x* and January 21, 2020 was recorded as the time “1,” the growth speed (*y′*) and time (*x*) were expressed as the following function:

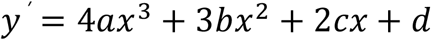

By drawing the derivative function of the fitted curves, we found that changes of the growth speed (*y′*) were accorded with our hypothesis. The growth speed (*y′*) increases firstly and declines sequentially after reaching the Speed_max_.

### The plateau of cumulative confirmed patients of COVID-19

Before February 23, 2020, the growth speed basically reached zero (new confirmed patients≤1 per day) for most areas except Hubei. If one area reported zero new cases for at least two days, the corresponding number of cumulative confirmed patients with COVID-19 was defined as the plateau number. If an area did not meet the criterion of zero-growth speed for two days, then the number of cumulative confirmed patients on February 23, 2020 was recorded as the plateau number.

### Software, instruments and statistics

In this study, model fitting was implemented by IBM SPSS Statistics 23.0 and Microsoft Office Excel 2010. The fitted functions and correlation coefficients were calculated by IBM SPSS Statistics 23.0 and Microsoft Office Excel 2010 as well. The curve of the derivative function was created by the drawing instrument from http://www.zuotu.91maths.com/, and the point of Speed_max_ was calculated by this website as well. The heat map of China was generated by the drawing instrument from http://www.dituhui.com/.

## Results

### Model fitting for known data in mainland China

First, we performed model fitting of different areas in mainland China except for Xizang and Qinghai because the confirmed number of cases were too few in these areas to model. The fitted curves were good, and all correlation coefficients R^2^ > 0.99 except for those in Jilin (R^2^ = 0.989) and Hubei (R^2^ = 0.988). Areas were divided into three parts: areas adjoined to Hubei (Figure 1), northern areas (Figure 2), and southern areas (Figure 3) of China. Because of an adjustment to confirmation criteria, the number of confirmed patients in Wuhan surged on February 22, 2020. Similarly, 200 new confirmed patients surged in Shandong on February 19, 2020 due to an outbreak of COVID-19 in a prison. However, the growth speed had basically reached zero since February 17, 2020 in Shandong. To eliminate the influence of this specific incident and perform model fitting, we fit the model for Shandong with adjusted data.

**Figure 1.**
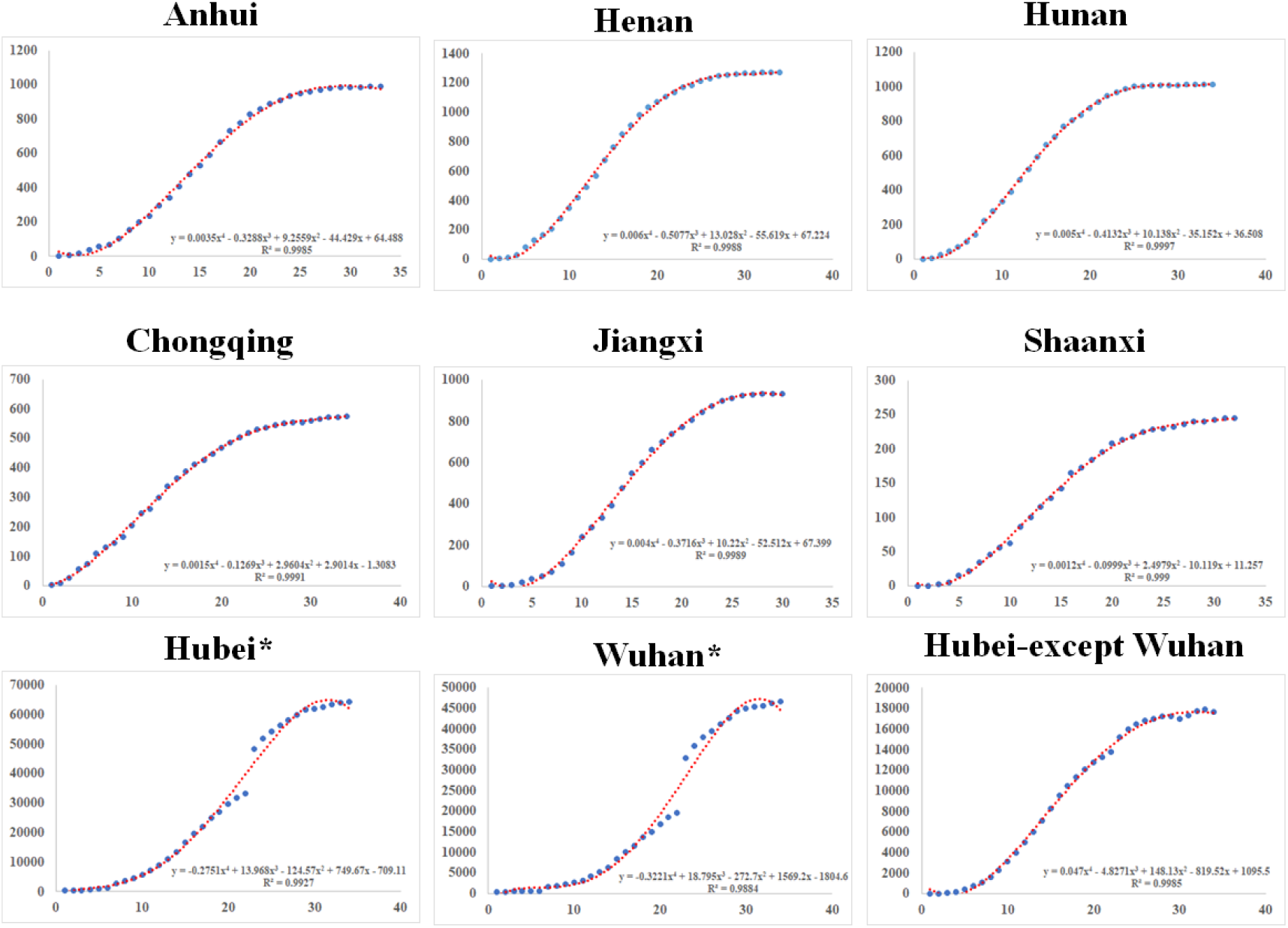
Model fitting in Hubei, Wuhan, Hubei-except Wuhan, and areas adjoined to Hubei. *Because the criterion of a confirmed case was adjusted, the number of confirmed patients in Wuhan (12,364) and Hubei (13,332) surged on February 22, 2020.

**Figure 2.**
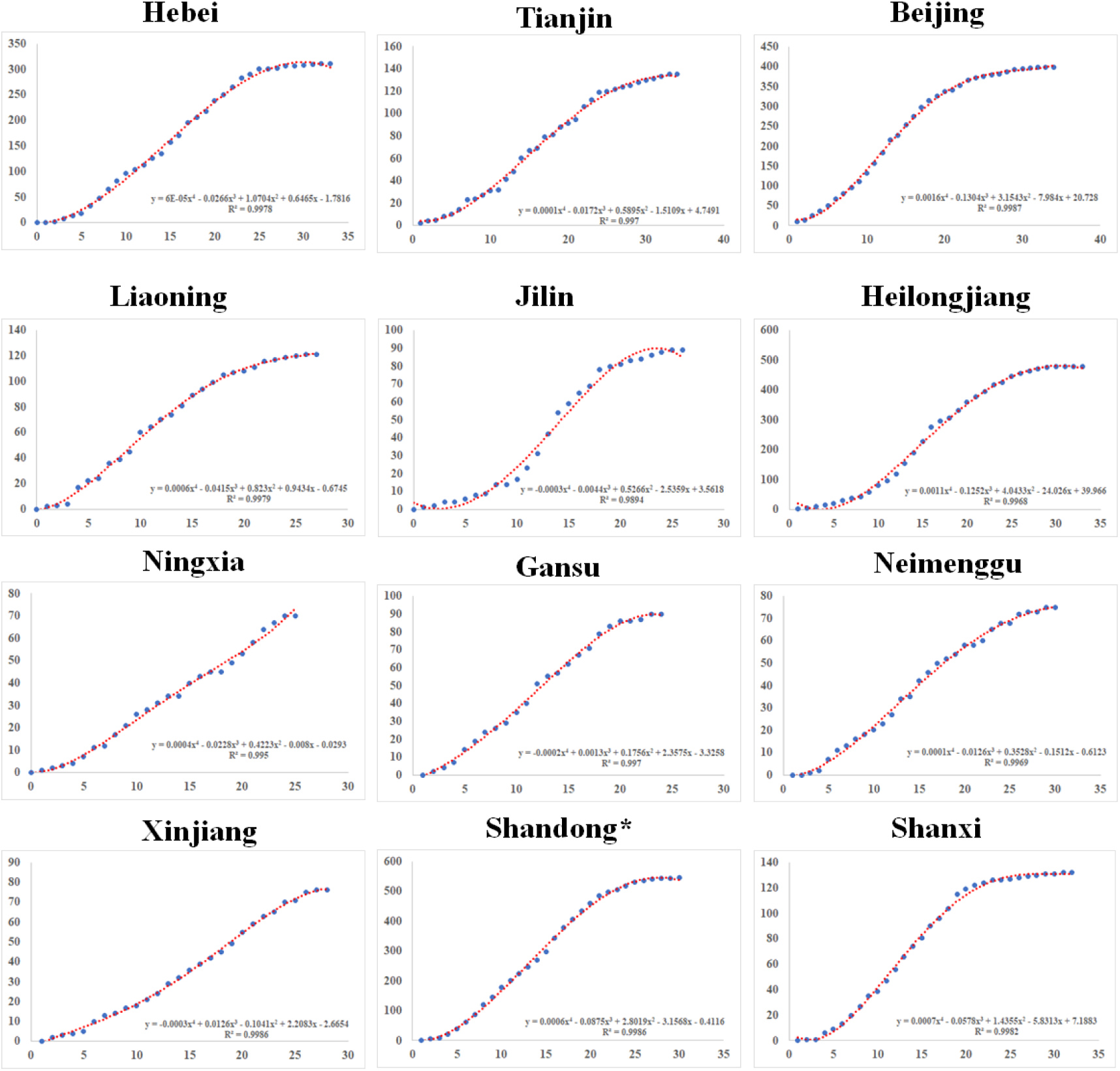
Model fitting in the northern area of mainland China. *A crack was generated in February 19, 2020 due to the surge of COVID-19 in a prison. To eliminate the influence of this specific incident, we performed model fitting with adjusted data for Shandong by removing the data of the prison outbreak.

**Figure 3.**
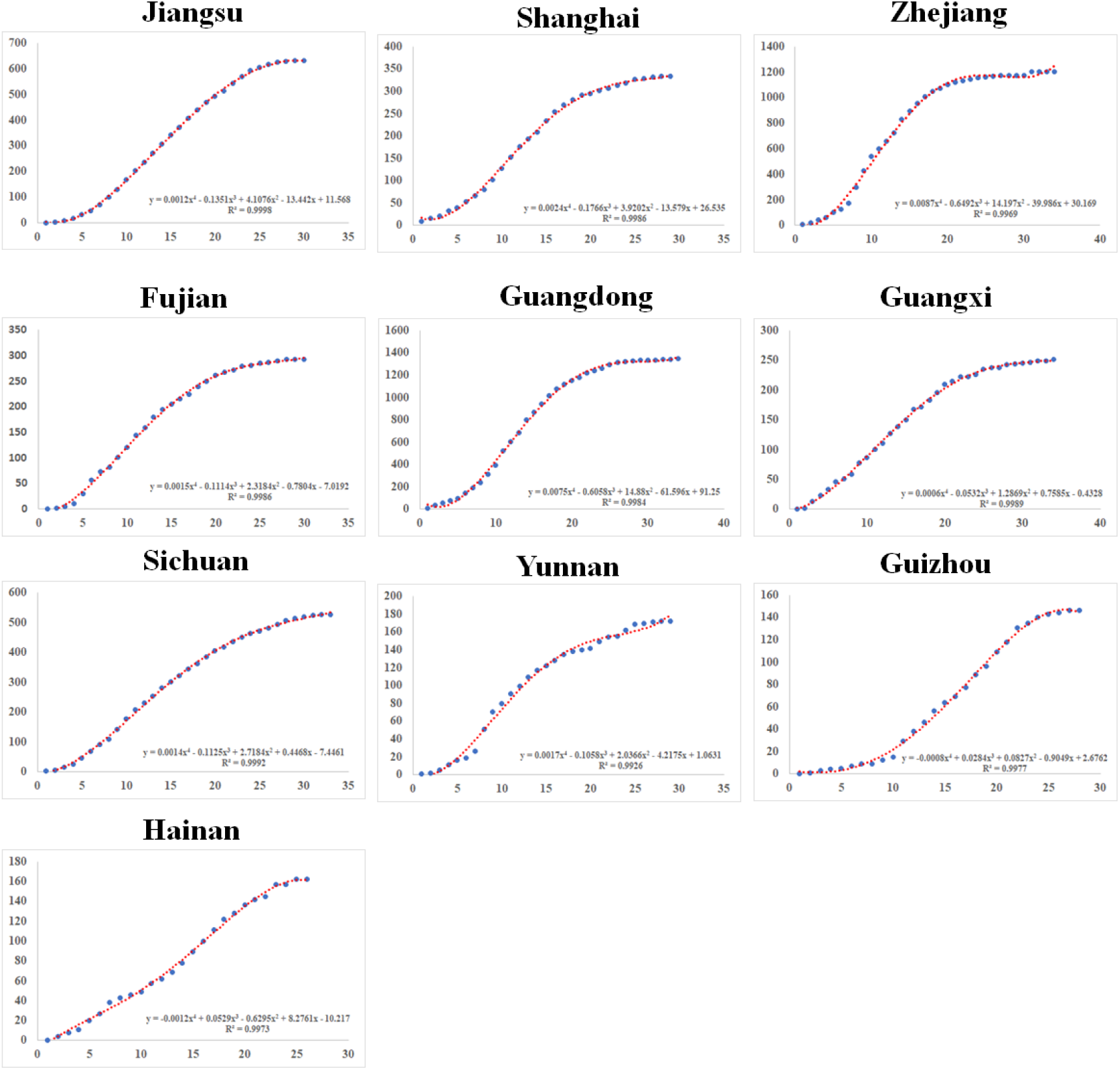
Model fitting in the southern area of mainland China.

### Calculation of Speed_max_

In this study, HongKong, Macao and Taiwan were not analyzed. Xizang, Xinjiang, Qinghai and Ningxia were not appropriate for Speed_max_ analyses because the confirmed patients were few or the growth speed was too smooth. Almost all areas in mainland China reached Speed_max_ within 14 days except Hebei, Heilongjiang, Hainan, Guizhou, and Hubei (Figure 4). Thus, the growth speed of confirmed cases began to decline in almost all areas before February 3, 2020. Because the outbreak of COVID-19 originated from Wuhan, Hubei Province, China, more time was required for Hubei and Wuhan to reach Speed_max_ (21–22 days and 23–24 days, respectively), although strict strategies were implemented in these regions.

**Figure 4.**
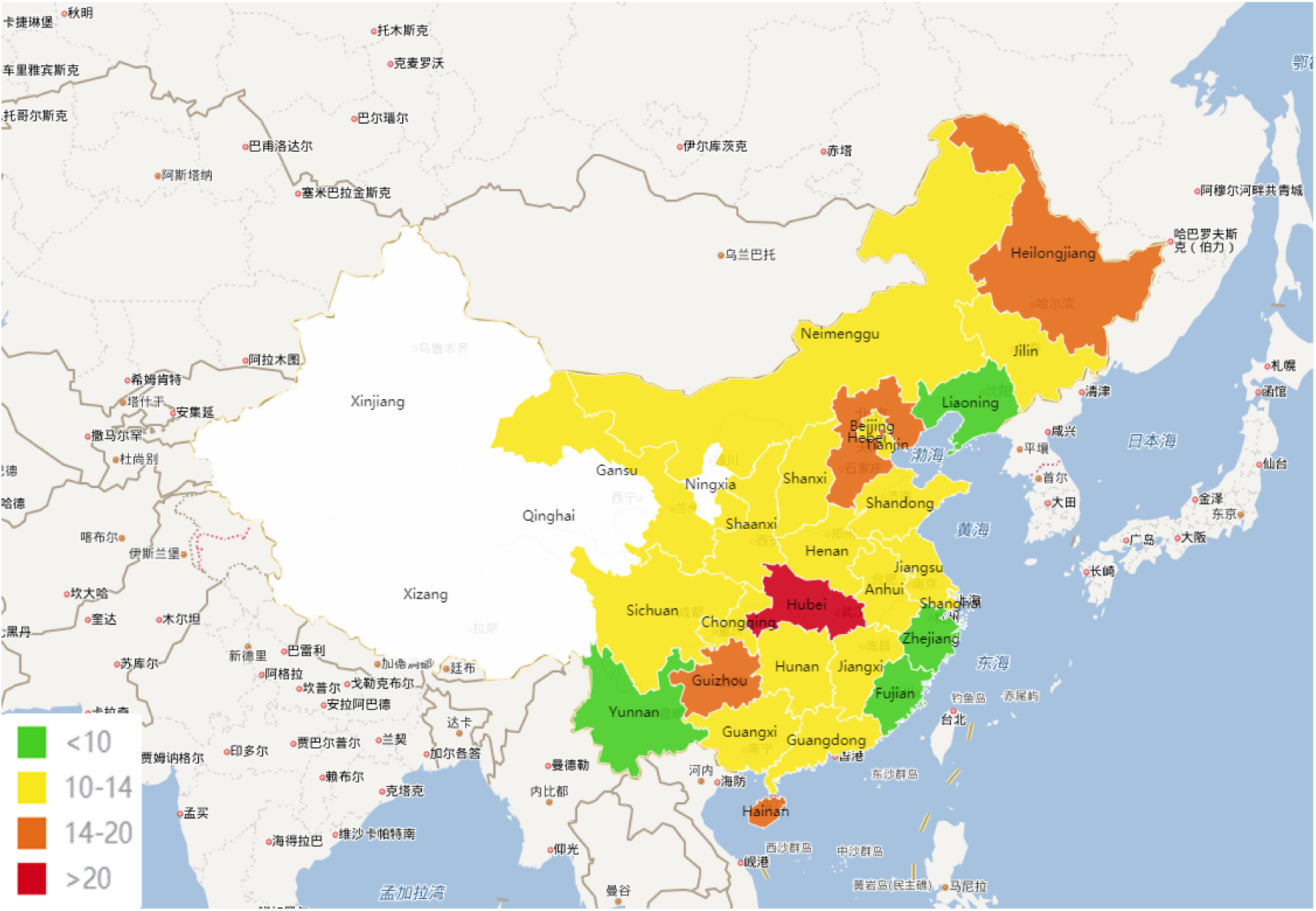
Heat map of the times to Speed_max_ in mainland China. *Xizang, Xinjiang, Qinghai, and Ningxia were not used for growth speed analyses because the numbers of confirmed patients were too few or the growth speed was too smooth.

The heat map of times for Speed_max_ suggests that the distance from other areas to Hubei have little influence arriving at Speed_max_ (Figure 4). After implementation of strict intervention strategies, the diffusion and deterioration of COVID-19 was inhibited quickly and effectively over China. Even the speed in Hubei, with the exception of Wuhan, reached Speed_max_ and began to decline in 14–15 days, which showed similar trends to those of other areas in mainland China.

As shown in Table 1, the average time to Speed_max_ was 11–12 days in China, with the exception of Hubei. Because little information was acquired at the beginning of the outbreak of COVID-19, it is understandable that Wuhan needed a longer time to reach the Speed_max_ point.

**Table 1.** Comparisons of the time from the first patient confirmed to Speed_max_ in different areas.

| Area* | Days from first patient confirmed to Speed<sub>max</sub> | Days from January 21, 2020 to Speed<sub>max</sub> |
| --- | --- | --- |
| China except Hubei* | 11.2 | 11.7 |
| Hubei except Wuhan* | / | 14 |
| Wuhan* | 63 | 23 |
| Hubei* | 61 | 21 |

\*In this study, Hong Kong, Macao and Taiwan were not analyzed. Xizang, Xinjiang, Qinghai, and Ningxia were not appropriate for analyses because the number of confirmed cases was few or the growth speed was too smooth. The time in China except Hubei was calculated by averaging the time of Speed<sub>max</sub> in other areas. The date of the first patient confirmed in Hubei except Wuhan was unknown. The date of first patient confirmed with unknown atypical pneumonia in Wuhan was recorded as December 8, 2019.

The detailed data for the time to Speed_max_ and emigration index was shown in Supplementary Table 1.

### The plateau of cumulative confirmed patients of COVID-19

The heat map of the plateau in mainland China (Figure 5) revealed that the plateau numbers were higher in areas adjoined to Hubei and southern areas of China and were lower in the northern area, especially in the western part of the northern area of China. After regression analysis and model-fitting, results suggested that the number of plateaus was significantly correlated with the emigration index, and the correlation coefficient reached 0.74 (Figure 6). However, the plateau numbers had no significant correlation with the time of Speed_max_ when compared with different areas in China. In Zhejiang, though the plateau numbers ranked third among mainland China except Hubei, the growth speed reached the Speed_max_ point and declined earlier than in many areas. This suggested that the spread of COVID-19 was controlled more quickly and effectively in Zhejiang. More detail about the number of plateaus is shown in Supplementary Table 1.

**Figure 5.**
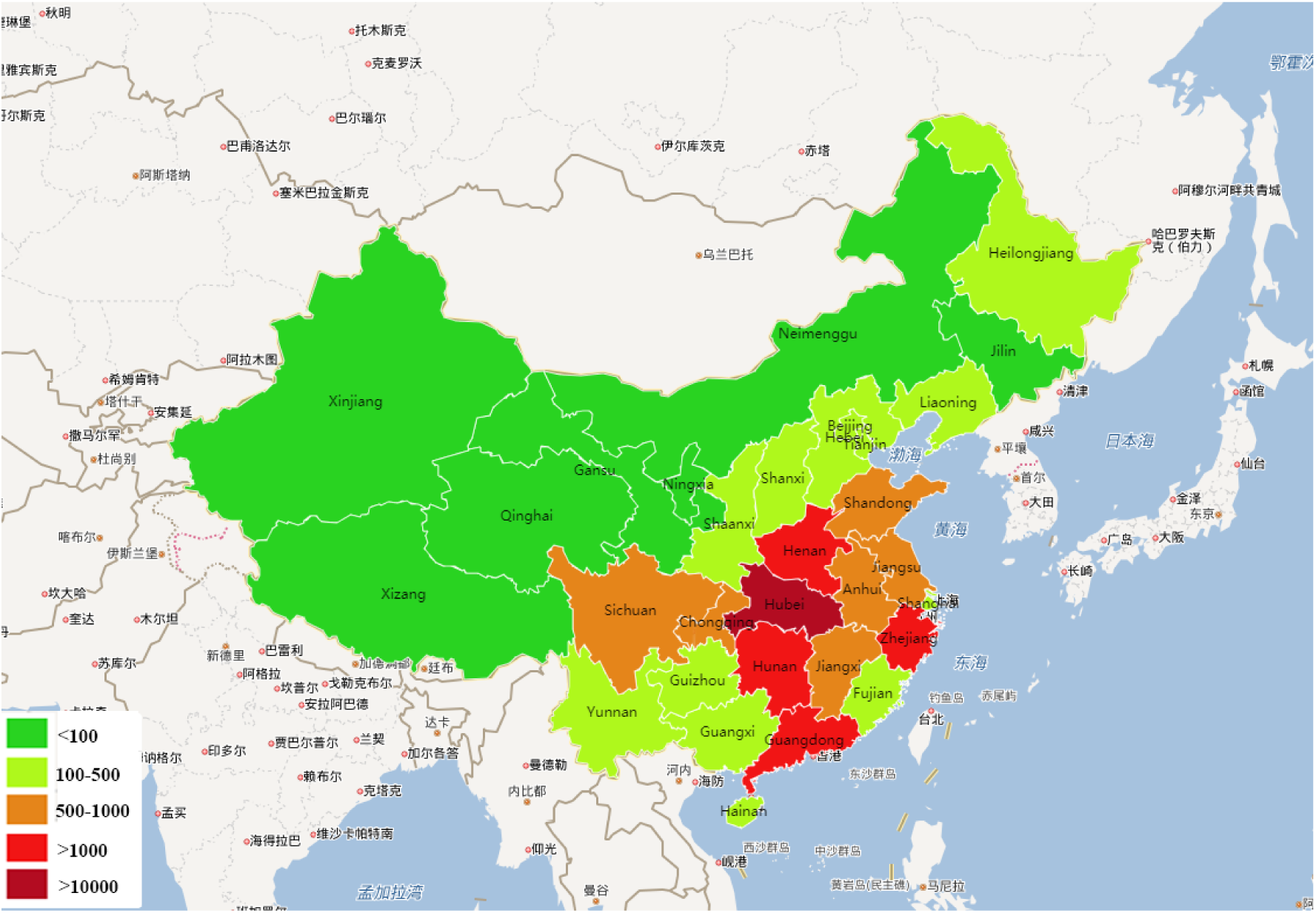
Heat map of the plateau number in mainland China.

**Figure 6.**
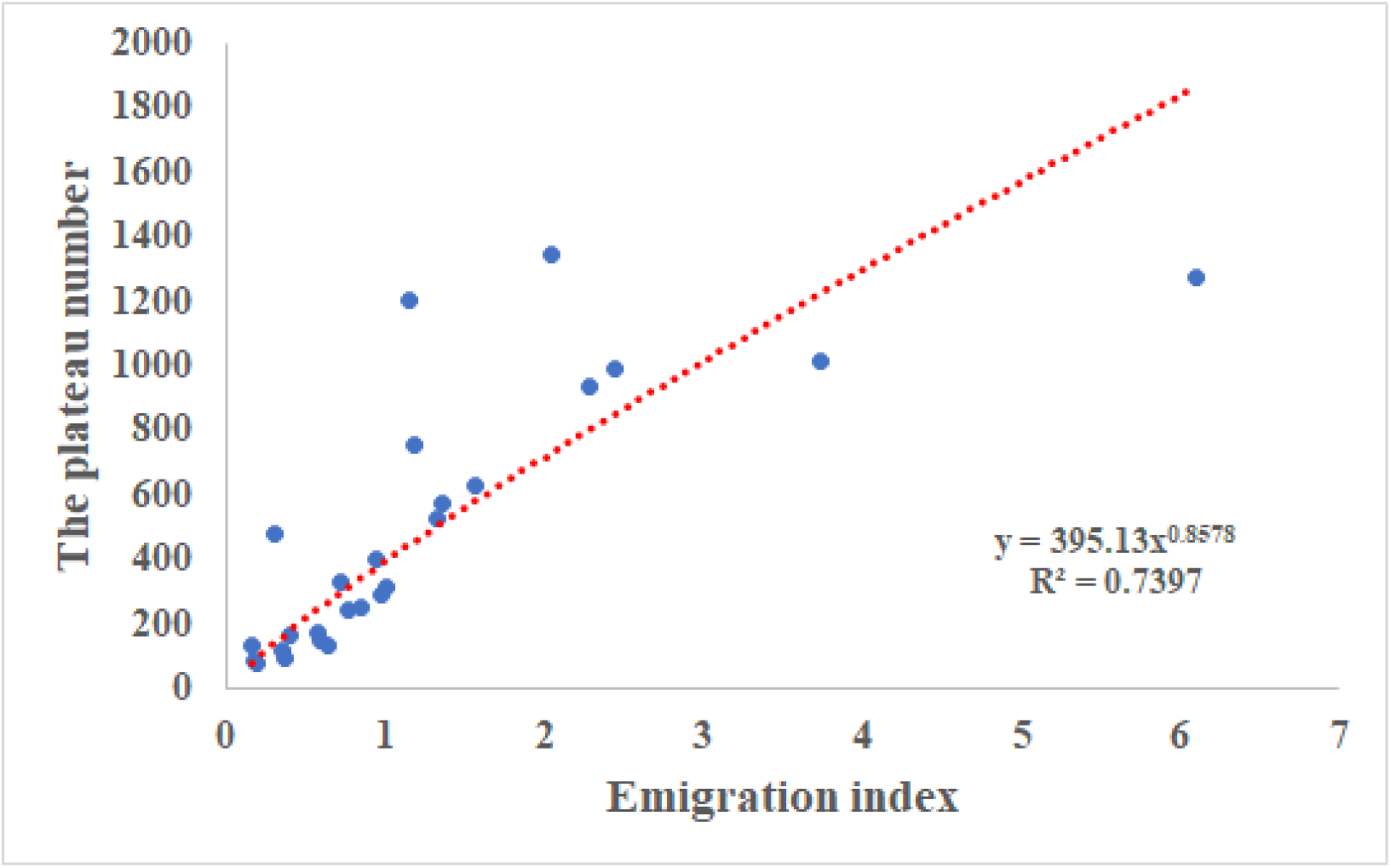
Correlation between emigration index and the plateau number.

## Discussion

COVID-19, a new and severe epidemic, has threatened public health and economic development in China ^[7]^. As of March 6, 2020, the number of cumulative confirmed cases of COVID-19 has reached 100,000 worldwide, and some countries such as Korea, Japan, Italy, and Iran are also suffering from the rapid spread of COVID-19 ^[4, 7, 8]^. The need for greater attention and intervention strategies to curb the spread of COVID-19 around the world is of paramount importance.

To inhibit the spread of COVID-19 as quickly as possible, the Chinese government took actions rapidly and implemented a series of strategies ^[5, 6, 9]^. For example, (1) President Xi of China conducted and deployed the control of COVID-19 personally, and all-of-government and all-of-society approaches were employed to contain viral spread; (2) Wuhan and other areas in Hubei were thoroughly quarantined, and the flow of people was completely restricted over the China; (3) a large number of doctors and nurses over China were assigned to Hubei to cure patients with COVID-19; (4) new special hospitals and vast mobile cabin hospitals were constructed quickly for accommodating patients with confirmed cases of COVID-19 and grading treatments; (5) critical medical resources and goods were regulated and distributed by the government; (6) traditional Chinese medicine and western medicine were combined to treat patients.

To better understand and estimate the effects of Chinese strategies on controlling he diffusion and deterioration of COVID-19 in mainland China, we implemented model fitting and designed an index named Speed_max_, which represented the time when the growth speed of the cumulative confirmed cases reached its maximum. According to our analyses, the growth speed generally increased first and decreased to zero quickly after reaching the Speed_max_ point. The area under the curve (AUC) of the derivative function represents the cumulative number of confirmed cases. Thus, per region, a shorter time to Speed_max_ generally indicates a smaller AUC, representing fewer cumulative confirmed cases when reaching the plateau. Therefore, to some extent, the time to Speed_max_ demonstrates the effectiveness of strategies on inhibiting the spread of virus. Notably, since the plateau number is affected by multiple factors, it is inappropriate to conclude that one area with shorter Speed_max_ means fewer plateau number than another area with longer Speed_max_. The Speed_max_ is useful for assessing changes of the plateau number itself.

According to our study, we found that these Chinese strategies for controlling the diffusion of COVID-19 are significantly effective and powerful. By implementing multiple Chinese strategies, the growth speed of confirmed patients reached Speed_max_ and began to decline within 14 days in almost all areas of mainland China. Yang *et al*. ^[9]^ also reported that a five-day delay in the implementation of the Chinese strategies would have increased the epidemic size in mainland China threefold. In addition, we found that the distance from other areas to Hubei and the number of plateaus have little influence arriving at Speed_max_. Once strict intervention strategies were implemented, the diffusion and deterioration of COVID-19 were inhibited quickly and effectively over China. With the exception of Wuhan, even Hubei, where the cumulative confirmed patients reached 17,680 on February 23, 2020, the speed of growth still reached Speed_max_ and began to decline in 14–15 days.

The present study has certain limitations. In this study, we simplified the model analysis and performed model fitting with known data. Although the classical susceptible-infectious-removed (SIR) and susceptible-exposed-infectious-removed (SEIR) models were not adopted, the quartic model showed a good fit to known data. This helps us to learn the trends of cumulative confirmed cases quickly and easily. However, our fitted models were all based on known data as it was more appropriate than forecasting future data trends. In addition, the changes in trends of basic reproduction number (R0) are not discussed in our study. The effects of Chinese strategies will be better understood if the time to Speed_max_ and changes in R0 are discussed together. Third, more data of different infectious diseases and studies are needed to verify the effectiveness and value of the index Speed_max_.

In conclusion, by analyzing the time to Speed_max_, our study suggests that Chinese strategies are highly effective on controlling the diffusion and deterioration of the Novel Coronavirus–Infected Pneumonia. These strategies therefore provide experience and effective guidelines for other countries to control the diffusion of COVID-19.

## Data Availability

All data has been supplied in the manuscript.

## Supplementary information

Supplementary Table 1

## Acknowledgments

None

